# Predicting Prostate Cancer Without a Prostate: A Potential Problem with AI

**DOI:** 10.1101/2025.03.04.25323311

**Authors:** Destie Provenzano, Murray Loew, Yuan James Rao, Vivek Batheja, Shawn Haji-Momenian

**Affiliations:** George Washington University School of Engineering and Applied Science. Washington, District of Columbia, USA; George Washington University School of Medicine and Health Sciences. Washington, District of Columbia, USA; Georgetown University School of Medicine and Health Sciences. Washington, District of Columbia, USA

**Author notes:** **Corresponding Author:** Dr. Shawn Haji-Momenian, Address: 3900 Reservoir Rd NW, Washington, DC 20007.

**Keywords:** Machine Learning, Prostate Cancer, Explainability, XAI, ResNet

## Abstract

Machine learning (ML) algorithms have demonstrated great potential for the identification and classification of prostate cancer from Magnetic Resonance (MR) Imaging data. Many of these algorithms remain a “black-box,” however, and debate persists as to how and if they should be explained. This study hypothesized that a widely-used family of methods, Convolutional Neural Networks (CNNs), may identify patterns that are not relevant to a clinician without model explainability. The purpose of this study was to determine if a CNN could classify prostate cancer on MR images without using the cancerous lesions -- or even the entire prostate – in the training process. We used 126 T2-weighted MR images each containing an abnormal prostate lesion to create two pairs of image sets: 1a) full cross-sectional images of the pelvis (full-CSI), 1b) full-CSI with prostate removed, 2a) segmented images of the prostate, and 2b) segmented images of the prostate with the lesion of interest removed. Residual Neural Network (ResNet) algorithms were trained and tested on the images, and accuracy and area under the receiver operating characteristic (AUC) were calculated. All algorithms performed well (accuracy of 81-99%, AUC of 0.83-0.99) even when a) trained on images containing the prostate/prostate lesion and tested on images with no prostate or prostate lesion or b) trained and tested on images with no prostate or prostate lesion. These findings support the need for explainable artificial intelligence (XAI) to ensure algorithms are arriving at clinically useful decisions.

**Significance Statement:** This study found that machine learning models built to classify clinically significant prostate cancer using predictive frameworks that rely on automatic feature detection (CNN - ResNet) can achieve high accuracy without evaluating the region of interest (in this case the prostate tissue). Although interesting, these models would not be viable in clinical practice. These results suggest rigorous testing and incorporation of explainability methods is urgently needed in machine learning models for clinical medicine to ensure models relying on automatic feature detection methods like CNNs select features that are clinically relevant.

## 1.0 Introduction

Prostate MRI for cancer detection is a promising area for the application of machine learning algorithms. ^1 2^ Many studies suggest that these algorithms are able to predict the presence and severity of prostate cancer on magnetic resonance (MR) images with high accuracy. ^3 4 5 6^ Algorithms that rely on Convolutional Neural Networks (CNNs) are considered revolutionary due to their ability to automatically create and identify many image features through a convolutional layer without human input. But the decision pathway used by these algorithms to find a suitable predictive pattern lack inherent explainability. Several models have been developed to “work backwards” to identify the image features most critical to the algorithm, including SHAP^7^, LIME^8^, and Grad-CAM^9^. But even these models have inherent shortcomings. ^10^ Furthermore, there remains debate as to how these machine learning (ML) models could be explained, what is required to ensure clinical usability in the explanation, and if these predictive models should be explained at all.^11^

We recently developed a Residual Neural Network (ResNet) that was able to accurately predict clinically significant prostate cancer (csPCA) from a single-institutional prostate MR image dataset.^12^ Further assessment of this algorithm’s most highly weighted feature maps (which are the most important pixel subsets used by the algorithm to “interpret” the image) showed the algorithm to be using image data outside the prostate lesion of interest. The purpose of this study is to determine if a ML algorithm can identify the presence of prostate cancer on MR images that omit the cancerous lesions or even the entire prostate. Such findings would have significant implications for the trustworthiness of these algorithms and suggest the need for better explainable artificial intelligence (XAI) to ensure that medical algorithms are using pertinent inputs to arrive at critical diagnostic outputs.

## 2. Results

### 2.1 Model Results

Models that were trained and tested on images containing the prostate and prostate lesion (i.e., full CSI and segmented prostate) were highly accurate (94%, p < 0.05) in the identification of csPCA. Models trained and tested on images without the prostate and prostate lesion (i.e., full CSI-without-prostate and segmented prostate-without-lesion) were also very accurate (90-99%, p<0.05). Algorithm accuracy decreased but remained relatively high (83-89%, p < 0.05) when trained on images with the prostate and prostate lesion (i.e., full CSI and segmented prostate) but tested on images without the prostate or prostate lesion (i.e., full CSI-without-prostate and segmented prostate-without-lesion). Accuracy remained very high (90-99%, p < 0.05) when training and testing on images without the prostate or prostate lesion (i.e., full CSI-without-prostate and segmented prostate-without-lesion). All models were statistically significant by shuffle test at p < 0.01 and by t-test on 5-fold cross-validated average data at p < 0.05. Model accuracy and AUC results are summarized in table 1.

**Table 1:**
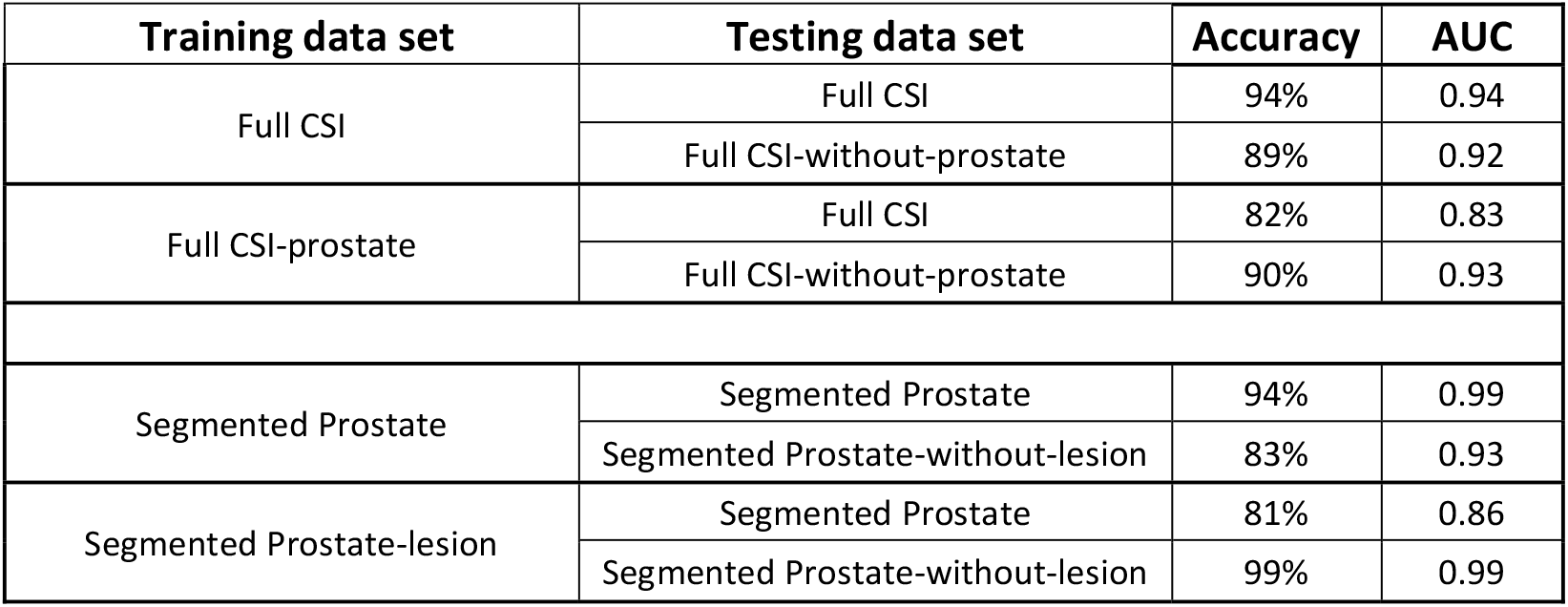
Model accuracy and AUC aggregated into averages across the 5-fold cross-validated datasets for each model and testing set combination for prediction of csPCA vs ncsL prostate lesion. 1. All model results were statistically significant at p < 0.01 by Shuffle Test. 2. All model results were statistically significant at p < 0.05 by t-test on the 5 cross-validated results for each final average.

| Training data set | Testing data set | Accuracy | AUC |
| --- | --- | --- | --- |
| Full CSI | Full CSI | 94% | 0.94 |
|  | Full CSI-without-prostate | 89% | 0.92 |
| Full CSI-prostate | Full CSI | 82% | 0.83 |
|  | Full CSI-without-prostate | 90% | 0.93 |
| Segmented Prostate | Segmented Prostate | 94% | 0.99 |
|  | Segmented Prostate-without-lesion | 83% | 0.93 |
| Segmented Prostate-lesion | Segmented Prostate | 81% | 0.86 |
|  | Segmented Prostate-without-lesion | 99% | 0.99 |

## 3.0 Discussion

Machine learning algorithms may be able to add great value and speed in the screening and interpretation of medical images.^13^ These algorithms are showing great potential in their diagnostic accuracy.^14^ The results of this study demonstrate that while ML algorithms appear to be highly accurate in their prediction of csPCA, they appear to be using non-clinically relevant image features to arrive at their prediction.

The algorithm that was trained and tested on the full CSI data had high accuracy (94%). These results are concordant with other published models that have achieved high accuracies (>90%) for prediction of prostate cancer on T2W images.^15^ These results appear highly promising as it appears the algorithm is correctly interpreting the imaging to arrive at the appropriate classification. The algorithm trained on full CSI data but tested on full CSI data-without-prostate (i.e., prostate removed) continued to have relatively high accuracy (89%). To maintain such high accuracy, the algorithm must be using imaging features from structures outside the prostate (periprostatic fat, rectum, pelvic musculature, bony pelvis) to determine if there is csPCA inside the absent gland. This is further confirmed by the continued high accuracy (90%) of training and testing on full CSI-without-prostate data. The gland containing the lesion of interest is not needed for the algorithm to accurately classify the image. While technically unclear due to the black-box nature of these models, the algorithm has found an incidental pattern of features in these extraneous structures that are predictive of the presence of csPCA in the gland. It is extremely unlikely that those structures have changes that can indicate the presence of cancer in the prostate. Even if there were theoretical cellular/molecular changes in these structures as a result of the presence or absence of prostate cancer, it would remain extremely unlikely that these changes would result in macroscopic image findings that are detectable by MR imaging.

Similar testing was performed on image data containing the segmented prostate gland to determine if the algorithm would become more reliant on the lesion of interest once large amounts of extraneous image data were removed. The algorithm had high accuracy (94%) when trained and tested on segmented prostate images. Accuracy decreased but remained relatively high (83%) when trained on segmented prostate but tested on segmented prostate--without-lesion. Accuracy was near perfect (99%) when training and testing on segmented prostate-without-lesion data. Even within the smaller segmented image that contained only the prostate, the algorithm was using features outside the lesion of interest to determine classification. These findings strongly suggest that the high performance of the algorithms on image sets containing the prostate and prostate lesion are also likely based on the use of image data outside the prostate and/or lesion of interest.

Radiologists use the well-established “Prostate Imaging – Reporting and Data System”^16^ guidelines in their interpretation of the likelihood of csPCA in prostate lesions. ML models such as the ResNet model used in this study utilize a convolutional layer to look for patterns anywhere in the image; this study demonstrated that these patterns can be using irrelevant inputs given the omission of the organ/lesion of interest. This raises significant questions about these models: are these models truly useful and trustworthy when it appears that they are using nonsensical inputs (periprostatic tissues or extra-lesional intra-prostatic tissue) to determine a critical output (presence of csPCA)? Further inclusion of algorithm explainability will likely be warranted with publication of promising AI data. Jin et al. published a set of five requirements to assess the efficacy and generalizability of algorithms’ performance, including understandable XAI metrics.^17^ Inclusion of some form of model explainability will also be a challenging task; there are currently many XAI techniques, each with its own strengths and weaknesses. ^18 19^ Most current model explainability techniques are qualitative, which can lead to interpreters incorrectly assuming features are important and to a gap between a model developer’s need for feature importance and a clinician’s need for features to be clinically relevant. ^20 21 22^

This study has some limitations. The study consists of a relatively small homogeneous patient population from a single institution. It is possible that a larger multi-institutional more heterogeneous image dataset could force the algorithm to become more reliant on the lesion of interest, although such studies are difficult to conduct, as illustrated by the paucity of such studies in the literature.^23^ Further segmentation of the prostate according to zonal anatomy could better focus the algorithm on the lesion of interest, although further zonal segmentation requires more work and reduces the efficiencies of algorithms using simple image inputs. Additionally, the ground truth in this study was established by MR-guided prostate biopsy. Biopsies can yield false negatives, placing limitations on our ground truth. Furthermore, it is theoretically possible that the algorithm correctly identified csPCA elsewhere in the prostate gland when it classified ncsL images as cancerous (radiology-whole-mount pathology correlation was not possible in this study and would require a prostatectomy cohort). This scenario remains very unlikely, however, as prostate MRI has a high negative predictive value for exclusion of csPCA^24^; it would therefore be very unlikely for cancer to be present elsewhere on the image slice (and images with more than one lesion of interest were excluded).

## 4.0 Methods

### 4.1 Data Collection and Preparation

Prostate MR image data from the ProstateX dataset on The Cancer Imaging Archive (TCIA)^25^ was used to develop a series of predictive models for this study. ProstateX consists of 330 multi-parametric prostate MRIs from Radboud University Medical Center (RUMC) collected on Siemens MAGNETOM Trio and Skyra 3T Magnetic Resonance (MR) scanners. Radiologists at RUMC interpreted the MRI exams and identified abnormal lesions warranting biopsy; the centroid of the lesions are provided as part of this dataset. The small field of view axial T2-weighted turbo spin echo sequence of the lower pelvis containing the lesion centroid was used for this study, with slice thickness of 3.6 mm and in-plane resolution of 0.5 mm. Gleason score was confirmed by prostate biopsy at RUMC. For the purposes of this study, “clinically significant” prostate cancer (csPCA) was defined as Gleason Score ≥ 3+4 (Gleason Grade ≥ 2) and “non-clinically significant” lesions (ncsL) were defined as Gleason score ≤ 3+3 (Gleason grade of ≤ 1).^26^

A balanced subset of image data from ProstateX was created by using 63 csPCA and 63 ncsL full cross-sectional images (full CSI). Each image contained only one prostate lesion per slice; images with more than one lesion per slice were excluded. A radiologist experienced in prostate MRI at our institution (S.H.M.) segmented the prostate on all 126 full CSIs to generate masks. The masks were used to generate two additional sets of image data: a) full cross-sectional images of the pelvis minus the prostate (full CSI-without-prostate) and b) segmented image of the prostate (segmented prostate). The prostate lesion centroid was then used to remove the lesion of interest from the segmented prostate images. A square mask of fixed size was used on all images to subtract the lesion of interest to produce the segmented prostate--without-lesion. The size of this square was 96 x 96 pixels and was determined by calculating the length of the largest lesion of interest and adding a “buffer” zone equivalent to the pixel size of a final ResNet generated feature. These two pairs of image sets (full CSI and full CSI-without-prostate, and segmented prostate and segmented prostate-without-lesion) were then used for training and testing purposes, as shown in Figure 1.

**Figure 1.**
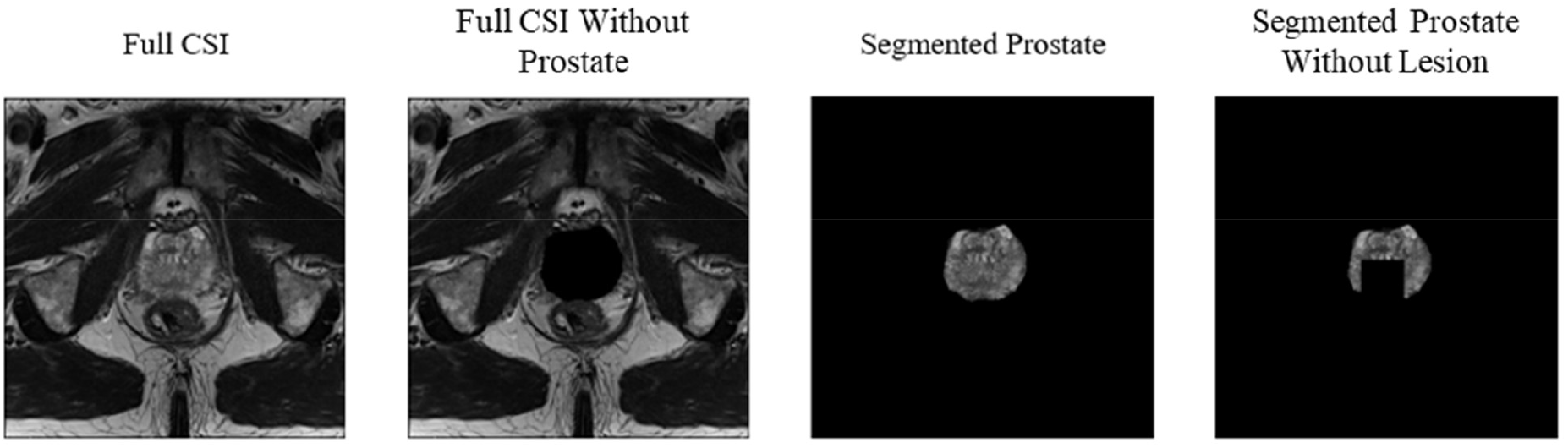
Example images used for model training of the four groupings of prostate images.

### 4.2 Model Training and Testing

Four Residual Neural Network (ResNet) models were trained on 1a) full CSI and 1b) full CSI-without-prostate, 2a) segmented prostate and 2b) segmented prostate-without-lesion image data to predict the presence of clinically-significant prostate cancer. We used the Python Tensorflow package on 80/20 training/testing splits of the two classes. Testing was performed on both image types within each pair, which tested the removal of the prostate or prostate lesion. Training and testing sets are summarized in Table 1. A transfer learning process in which all earlier model layers of the ResNet were held constant was employed using the ImageNet model weights.^27^ A binary sigmoid function mapped final image predictions to csPCA/ncsL status.

Model statistical significance was validated through 5-fold cross-validation using 5 folds of 80/20 training/testing splits. Accuracy and Area Under the Receiver Operating Characteristic ROC Curve (AUC) are reported for each model.^28^ A Randomization (“Shuffle”) Test was used at p < 0.05 to ensure significance of each final model, in which labels (csPCA/ncsL) were randomly assigned to each image to see if any shuffled run could achieve the same accuracy as the final model.^29^

## 5.0 Conclusion

This study demonstrated that machine learning algorithms are able to accurately predict the presence of csPCA on images that omit the prostate or prostate lesion. It also strongly suggests that their high performance on images containing a small field of view of the pelvis or segmented prostate is due to the utilization of image data outside the prostate and/or lesion of interest. The many layers of these neural networks are able to find incidental predictive patterns within nonsensical extraneous features. This raises significant concerns about the utility and trustworthiness of these algorithms. Further development (and inclusion) of machine learning explainability metrics and XAI are warranted to better explain the workings of these models to ensure that clinically reasonable inputs are utilized for such critical outputs.

## Data Availability

All data produced are available online at The Cancer Imaging Archive, ProstateX Collection. DOI: 10.7937/K9TCIA.2017.MURS5CL

https://www.cancerimagingarchive.net/collection/prostatex/

## Abbreviations

ML: Machine Learning
ResNet: Residual Neural Network
pCA: Prostate Cancer
csPCA: Clinically Significant Prostate Cancer
ncsL: Non-Clinically Significant Prostate Cancer
CSI: Cross Sectional Image
AUC: Area Under the Receiver Operator Characteristic Curve
XAI: Explainable Artificial Intelligence

## 6.0 Acknowledgments and funding sources

The authors of this study declare no outside funding.

## References

1. Baydoun, A., Jia, A.Y., Zaorsky, N.G. et al. Artificial intelligence applications in prostate cancer. Prostate Cancer Prostatic Dis 27, 37–45 (2024). 10.1038/s41391-023-00684-0

2. Turkbey B, Haider MA. Deep learning-based artificial intelligence applications in prostate MRI: brief summary. Br J Radiol. 2022 Mar 1;95(1131):20210563. doi: 10.1259/bjr.20210563. Epub 2021 Dec 3. PMID: 34860562; PMCID: PMC8978238.

3. Cuocolo R, Cipullo MB, Stanzione A, Romeo V, Green R, Cantoni V, Ponsiglione A, Ugga L, Imbriaco M. Machine learning for the identification of clinically significant prostate cancer on MRI: a meta-analysis. Eur Radiol. 2020 Dec;30(12):6877–6887. doi: 10.1007/s00330-020-07027-w. Epub 2020 Jun 30. PMID: 32607629.

4. Wildeboer RR, van Sloun RJG, Wijkstra H, Mischi M. Artificial intelligence in multiparametric prostate cancer imaging with focus on deep-learning methods. Comput Methods Programs Biomed. 2020 Jun;189:105316. doi: 10.1016/j.cmpb.2020.105316. Epub 2020 Jan 7. PMID: 31951873.

5. Riaz IB, Harmon S, Chen Z, Naqvi SAA, Cheng L. Applications of Artificial Intelligence in Prostate Cancer Care: A Path to Enhanced Efficiency and Outcomes. Am Soc Clin Oncol Educ Book. 2024 Jun;44(3):e438516. doi: 10.1200/EDBK_438516. PMID: 38935882.

6. Thomas M, Murali S, Simpson BSS, Freeman A, Kirkham A, Kelly D, Whitaker HC, Zhao Y, Emberton M, Norris JM. Use of artificial intelligence in the detection of primary prostate cancer in multiparametric MRI with its clinical outcomes: a protocol for a systematic review and meta-analysis. BMJ Open. 2023 Aug 22;13(8):e074009. doi: 10.1136/bmjopen-2023-074009. PMID: 37607794; PMCID: PMC10445392.

7. Lundberg, Scott. “A unified approach to interpreting model predictions.” arXiv preprint 1705.07874 (2017).

8. Ribeiro, M. T., Singh, S., & Guestrin, C. (2016). “Why Should I Trust You?”: Explaining the Predictions of Any Classifier (Version 3). arXiv. 10.48550/ARXIV.1602.04938

9. Selvaraju, R.R., Cogswell, M., Das, A. et al. Grad-CAM: Visual Explanations from Deep Networks via Gradient-Based Localization. Int J Comput Vis 128, 336–359 (2020). 10.1007/s11263-019-01228-7

10. Chaddad A, Peng J, Xu J, Bouridane A. Survey of Explainable AI Techniques in Healthcare. Sensors (Basel). 2023 Jan 5;23(2):634. doi: 10.3390/s23020634. PMID: 36679430; PMCID: PMC9862413.

11. Amann J, Blasimme A, Vayena E, Frey D, Madai VI; Precise4Q consortium. Explainability for artificial intelligence in healthcare: a multidisciplinary perspective. BMC Med Inform Decis Mak. 2020 Nov 30;20(1):310. doi: 10.1186/s12911-020-01332-6. PMID: 33256715; PMCID: PMC7706019.

12. Provenzano D, Melnyk O, Imtiaz D, McSweeney B, Nemirovsky D, Wynne M, Whalen M, Rao YJ, Loew M, Haji-Momenian S. Machine Learning Algorithm Accuracy Using Single-versus Multi-Institutional Image Data in the Classification of Prostate MRI Lesions. Applied Sciences. 2023; 13(2):1088. 10.3390/app13021088

13. Irbaz Bin Riaz et al., Applications of Artificial Intelligence in Prostate Cancer Care: A Path to Enhanced Efficiency and Outcomes. Am Soc Clin Oncol Educ Book 44, e438516(2024). DOI:10.1200/EDBK_438516

14. Castillo T JM, Arif M, Starmans MPA, Niessen WJ, Bangma CH, Schoots IG, Veenland JF. Classification of Clinically Significant Prostate Cancer on Multi-Parametric MRI: A Validation Study Comparing Deep Learning and Radiomics. Cancers (Basel). 2021 Dec 21;14(1):12. doi: 10.3390/cancers14010012. PMID: 35008177; PMCID: PMC8749796.

15. Li H, Lee CH, Chia D, Lin Z, Huang W, Tan CH. Machine Learning in Prostate MRI for Prostate Cancer: Current Status and Future Opportunities. Diagnostics (Basel). 2022 Jan 24;12(2):289. doi: 10.3390/diagnostics12020289. PMID: 35204380; PMCID: PMC8870978.

16. Turkbey B, Rosenkrantz AB, Haider MA, et al. Prostate Imaging Reporting and Data System Version 2.1: 2019 Update of Prostate Imaging Reporting and Data System Version 2. Eur Urol. 2019;76(3):340–351. doi:10.1016/j.eururo.2019.02.033

17. Weina Jin, Xiaoxiao Li, Mostafa Fatehi, Ghassan Hamarneh, Guidelines and evaluation of clinical explainable AI in medical image analysis, Medical Image Analysis, Volume 84, 2023, 102684, ISSN 1361-8415, 10.1016/j.media.2022.102684.

18. Nguyen H.T.T., Cao H.Q., Nguyen K.V.T., Pham N.D.K. Evaluation of Explainable Artificial Intelligence: SHAP, LIME, and CAM; Proceedings of the FPT AI Conference 2021; Ha Noi, Viet Nam. 6–7 May 2021; pp. 1–6.

19. Van der Velden B.H., Kuijf H.J., Gilhuijs K.G., Viergever M.A. Explainable artificial intelligence (XAI) in deep learning-based medical image analysis. Med. Image Anal. 2022;79:102470. doi: 10.1016/j.media.2022.102470.

20. Lopes P., Silva E., Braga C., Oliveira T., Rosado L. XAI Systems Evaluation: A Review of Human and Computer-Centred Methods. Appl. Sci. 2022;12:9423. doi: 10.3390/app12199423.

21. Ghassemi, M., Oakden-Rayner, L., & Beam, A. L. (2021). The false hope of current approaches to explainable artificial intelligence in health care. In The Lancet Digital Health (Vol. 3, Issue 11, pp. e745–e750). Elsevier BV. 10.1016/s2589-7500(21)00208-9

22. Bienefeld N, Boss JM, Lüthy R, Brodbeck D, Azzati J, Blaser M, Willms J, Keller E. Solving the explainable AI conundrum by bridging clinicians’ needs and developers’ goals. NPJ Digit Med. 2023 May 22;6(1):94. doi: 10.1038/s41746-023-00837-4. PMID: 37217779; PMCID: PMC10202353.

23. Teo ZL, Jin L, Li S, Miao D, Zhang X, Ng WY, Tan TF, Lee DM, Chua KJ, Heng J, Liu Y, Goh RSM, Ting DSW. Federated machine learning in healthcare: A systematic review on clinical applications and technical architecture. Cell Rep Med. 2024 Feb 20;5(2):101419. doi: 10.1016/j.xcrm.2024.101419. Epub 2024 Feb 9. Erratum in: Cell Rep Med. 2024 Mar 19;5(3):101481. doi: 10.1016/j.xcrm.2024.101481. PMID: 38340728; PMCID: PMC10897620.

24. Sathianathen NJ, Omer A, Harriss E, Davies L, Kasivisvanathan V, Punwani S, Moore CM, Kastner C, Barrett T, Van Den Bergh RC, Eddy BA, Gleeson F, Macpherson R, Bryant RJ, Catto JWF, Murphy DG, Hamdy FC, Ahmed HU, Lamb AD. Negative Predictive Value of Multiparametric Magnetic Resonance Imaging in the Detection of Clinically Significant Prostate Cancer in the Prostate Imaging Reporting and Data System Era: A Systematic Review and Meta-analysis. Eur Urol. 2020 Sep;78(3):402–414. doi: 10.1016/j.eururo.2020.03.048. Epub 2020 May 20. PMID: 32444265.

25. Armato SG 3rd, Huisman H, Drukker K, Hadjiiski L, Kirby JS, Petrick N, Redmond G, Giger ML, Cha K, Mamonov A, Kalpathy-Cramer J, Farahani K. PROSTATEx Challenges for computerized classification of prostate lesions from multiparametric magnetic resonance images. J Med Imaging (Bellingham). 2018 Oct;5(4):044501. doi: 10.1117/1.JMI.5.4.044501. Epub 2018 Nov 10. PMID: 30840739; PMCID: PMC6228312.

26. Gleason DF. Classification of prostatic carcinomas. Cancer Chemother Rep. 1966 Mar;50(3):125–8.

27. Deng J, Dong W, Socher R, Li L-J, Li K, Fei-Fei L. Imagenet: A large-scale hierarchical image database. In: 2009 IEEE conference on computer vision and pattern recognition. 2009. p. 248–55.

28. Bradley, Andrew P. “The use of the area under the ROC curve in the evaluation of machine learning algorithms.” Pattern recognition 30.7 (1997): 1145–1159.

29. Wood, M. (2018). How sure are we? Two approaches to statistical inference (Version 1). arXiv. 10.48550/ARXIV.1803.06214

